# Impact of a Smart Phone-Based Educational Video on Bowel Preparation and Colonoscopy Adherence: A Propensity Score Matched Analysis

**DOI:** 10.1101/2025.03.13.25323456

**Authors:** Kimberline Chew, Denise Galeano, Daniel Behin, Akash Kumar, Rohit Maheshwari, Lital Aliasi-Sinai, William Southern, Shitij Arora

**Affiliations:** Department of Internal Medicine, Division of Gastroenterology, Montefiore Medical Center, Bronx, NY, USA; Division of Gastroenterology, Montefiore Medical Center, Bronx, NY, USA; HealthiPeople Corp, San Francisco, CA, USA; Department of Medicine, Icahn School of Medicine at Mount Sinai Morningside, and West Hospital, New York, NY, USA; Division of Hospital Medicine, Montefiore Medical Center, Bronx, NY, USA; Director of Inpatient Digital Innovation, Montefiore Medical Center, Bronx, NY, USA

**Keywords:** Videos, Digital, Innovation

## Abstract

**Background and Aims:** Inadequate bowel preparation and high cancellation rates hinder the effectiveness of colonoscopy screening programs. We aimed to evaluate the impact of a novel, personalized video messaging platform on bowel preparation quality and colonoscopy adherence.

**Methods:** We conducted a before-after intervention study at a large urban academic medical center, comparing outcomes between a historical control group (April-September 2022) and an intervention group receiving personalized video messages (July-November 2023). Primary outcomes were inadequate bowel preparation and colonoscopy cancellation/rescheduling rates. Propensity score matching and multivariable logistic regression were used to adjust for potential confounders.

**Results:** Among 1,647 patients who underwent colonoscopy, inadequate bowel preparation occurred in 12.0% of the intervention group versus 20.0% of the historical control group in the propensity score matched analysis (p=0.041). In the larger cohort of 2,802 scheduled patients, the propensity score matched analysis showed a trend toward a 31% reduction in odds of cancellation or rescheduling associated with the educational video, although this did not reach statistical significance (OR 0.69, 95% CI 0.47-1.01, p=0.056). For the logistical regression model, the video was associated with a 42% reduction in odds of cancellation or rescheduling (adjusted OR 0.58, 95% CI 0.44-0.77, p<0.001).

**Conclusions:** Implementation of a personalized video messaging platform was associated with significant improvements in bowel preparation quality and substantial reductions in colonoscopy cancellations/rescheduling. This easily scalable, low-cost intervention has the potential to enhance the efficiency and effectiveness of colonoscopy screening programs.

## Introduction

Colorectal cancer (CRC) is a major public health issue, as it is the third most commonly diagnosed cancer and second leading cause of cancer deaths in the United States.^1^ Screening for CRC is critical for early detection and prevention of cancer progression, yet screening rates remain suboptimal with just 68.8% of adults adherent to current US Preventive Services Task Force guidelines.^2^ Adequate bowel preparation is essential for high-quality screening colonoscopies that reliably detect adenomatous polyps and CRC.^3^ However, many patients struggle to follow bowel preparation instructions, resulting in suboptimal preparations that reduce procedural efficacy, efficiency, and safety while increasing costs.^3,4^ Patient-reported barriers to adequate bowel preparation include unpleasant taste, large fluid volume, timing of dose, comprehension of instructions, embarrassment, anxiety, transportation limitations, and low self-efficacy.^5^

Digital health interventions, particularly those utilizing video messages, have shown promise in promoting health behaviors and improving patient outcomes across various health domains. For instance, video-based education has been found to be more effective than text-based instructions in enhancing patients’ knowledge, self-efficacy, and adherence to complex health behaviors.^6^ The efficacy of video interventions is attributed to their ability to engage multiple sensory channels, illustrate abstract concepts, and provide a more personalized and empathetic communication approach.^7^

Several studies have explored the impact of educational videos on bowel preparation quality and colonoscopy outcomes.^8,9^ For example, Prakash et al. provided patients with access to an online educational video via a password-protected website and found improved bowel preparation quality compared to standard instructions.^8^ However, the videos used in these studies were often lengthy (>10 minutes) and required patients to actively seek out and watch the content on specific platforms. In contrast, the widespread use of smartphones and text messaging offers an opportunity to proactively deliver brief, engaging, and easily accessible video content directly to patients.

Despite the potential advantages of mobile-based short video interventions, their efficacy in optimizing bowel preparation and colonoscopy outcomes has not been thoroughly investigated. To bridge this knowledge gap, we aimed to evaluate the effectiveness of a short educational video delivered via text messaging in improving bowel preparation quality and reducing colonoscopy cancellations and rescheduling. We hypothesized that patients receiving the short video intervention would have a lower likelihood of inadequate bowel preparation and a reduced rate of cancellations and rescheduling compared to those receiving standard care. The findings of this study could provide valuable insights into the use of short video messages as simple, scalable, and patient-centered approaches to optimize colonoscopy outcomes and ultimately enhance the efficacy of CRC screening programs.

## Methods

### Study Design and Setting

We conducted a before-after intervention study to evaluate the impact of a personalized video messaging platform, ’Montefiore Videoreach,’ on bowel preparation quality and colonoscopy cancellation/rescheduling rates. The study was performed at Montefiore Medical Center, a large, urban, academic medical center in Bronx, New York, USA. The study period was divided into two phases: a baseline phase from April to September 2022 (historical control group) and an intervention phase (intervention group) from July to November 2023. The study periods were chosen to allow for a sufficient sample size while minimizing potential seasonal variations. The historical control period (April-September 2022) and intervention period (July-November 2023) were selected to capture similar seasonal patterns in colonoscopy scheduling.

### Participants

The overall study population included patients aged 18 years or older who met criteria for colon cancer screening and were scheduled for an outpatient screening colonoscopy at Montefiore Medical Center during the study period. Patients were excluded if they had a history of colorectal cancer, inflammatory bowel disease, or previous colorectal surgery.

Two distinct study cohorts were derived from this overall population for the analysis of the primary and secondary outcomes. All patients were scheduled for screening colonoscopies, and none were interventional (Figure 1):

**Figure 1:**
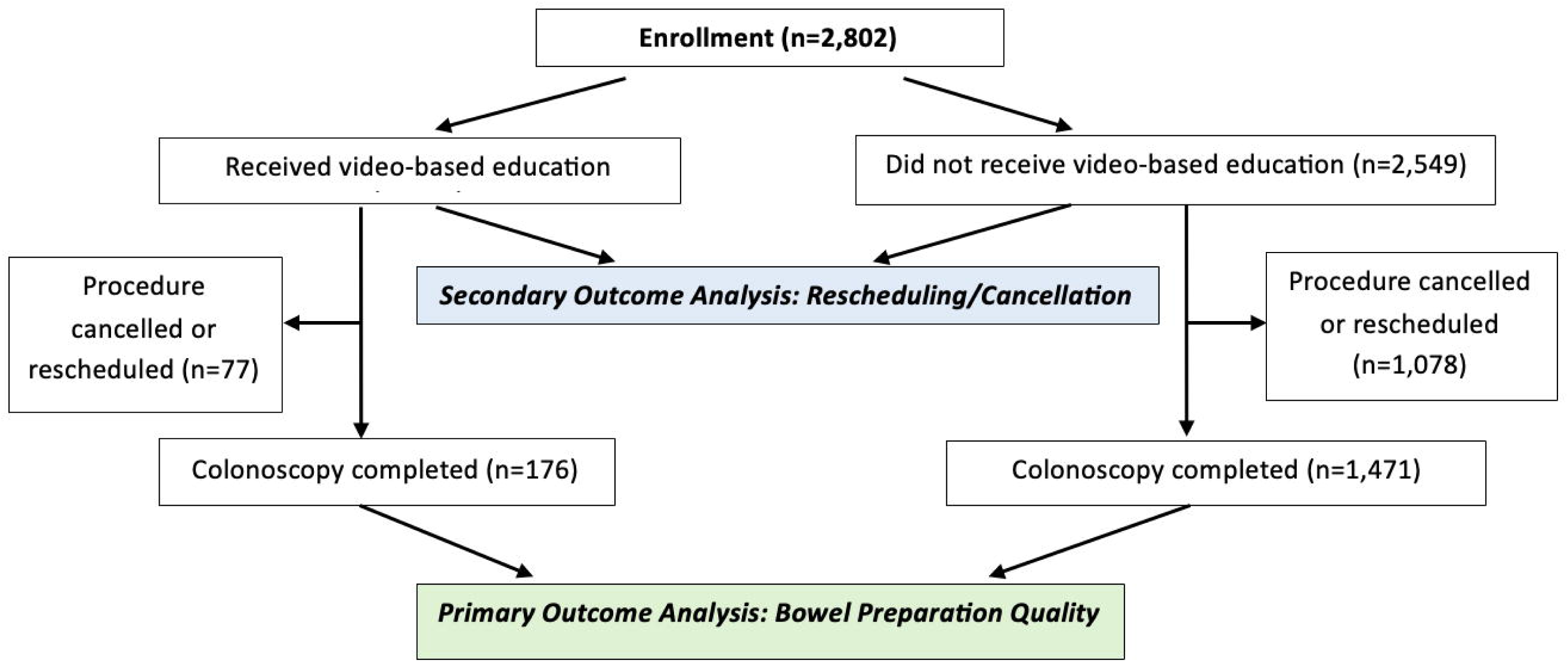
Patient Flow Chart for Primary and Secondary Cohorts

Primary outcome cohort: Patients who completed their scheduled outpatient screening colonoscopy at Montefiore Medical Center during the study period. This cohort was used to examine the association between the intervention video and inadequate bowel preparation.

Secondary outcome cohort: Patients who were scheduled for an outpatient screening colonoscopy at Montefiore Medical Center during the study period, regardless of whether they completed the procedure. This cohort was used to examine the association between the intervention video and the cancellation or rescheduling of the colonoscopy.

### Intervention (“Montefiore Videoreach”)

During the intervention phase, patients scheduled for a colonoscopy who agreed to participate in the study received a personalized educational video in addition to the standard written instructions. Patients were informed about the videos when called by our navigators to schedule their colonoscopies. Consent for the video was also obtained at that time. If the patient consented for the video, they were given information about how and when they would receive the videos. Depending on their preferred language, either English or Spanish, our navigators would allocate the appropriate video to the individual patient. The video, which was 90 seconds in duration, was sent via text message to patients’ smartphones starting at 1-day intervals in the 3 days before the scheduled procedure. The video provided a step-by-step guide to the 1-day Golytely bowel preparation protocol, including instructions on how to take the prescribed laxatives, dietary recommendations, and tips for staying hydrated. The video also emphasized the importance of proper bowel preparation for ensuring a high-quality colonoscopy and early detection of colorectal cancer. All patients scheduled for colonoscopy during the intervention phase were offered the educational video. Those who agreed to participate and provided a valid mobile phone number received the video messages.

### Outcomes

The primary outcome of the study was the quality of bowel preparation, which was assessed using the Boston Bowel Preparation Scale (BBPS) by the endoscopist performing the colonoscopy. The BBPS is a validated measure of bowel preparation quality, with scores ranging from 0 to 9 based on the cleanliness of the right, transverse, and left colon segments. A total BBPS score of ≥6 was considered adequate bowel preparation. The secondary outcome was the rate of colonoscopy cancellations and rescheduling, which was assessed by reviewing the electronic health record, where cancellations and rescheduling events were documented in a designated section.

### Data Collection

Demographic and clinical data, including age, gender, Charlson Comorbidity Index (calculated using ICD-10 codes input into calculator),^10^ insurance status, and preferred language, were collected from the electronic health record for both the primary and secondary cohorts. Bowel preparation quality and colonoscopy cancellation/rescheduling status were also obtained from the electronic health record.

### Statistical Analysis

Baseline characteristics, including age, sex, race/ethnicity, insurance status, Charlson comorbidity index, and primary language, were collected for all patients in both the historical control and intervention groups. These characteristics were compared between the two groups using t-tests for continuous variables and chi-square tests for categorical variables.

Propensity score matching was used to balance the distribution of potential confounders between the intervention and historical control groups. A logistic regression model was used to estimate the propensity score for each patient, which represented the probability of being in the intervention group based on the baseline characteristics.

Patients in the intervention group were then matched 1:1 with patients in the historical control group using a nearest-neighbor matching algorithm with a caliper width of 0.2 standard deviations.

The association between the educational video intervention and the outcomes (inadequate bowel preparation and colonoscopy cancellation/rescheduling) was assessed using logistic regression models. In the unmatched analyses, unadjusted and adjusted odds ratios (ORs) with 95% confidence intervals (CIs) were estimated. The adjusted models included age, sex, race/ethnicity, insurance status, Charlson comorbidity index, and primary language as covariates. In the propensity score matched analyses, conditional logistic regression was used to estimate the odds ratio for the association between the video intervention and the outcomes, accounting for the matched design.

All statistical analyses were performed using Stata version 17 (StataCorp, College Station, TX), and a two-sided p-value <0.05 was considered statistically significant.

## Results

### Study Population

The overall study population consisted of 2,802 patients scheduled for an outpatient screening colonoscopy during the study period.

For the primary outcome (inadequate bowel preparation), the unmatched study cohort included 1,647 patients who completed their colonoscopy, with 176 patients (10.7%) in the intervention group and 1,471 (89.3%) in the historical control group.

For the secondary outcome (colonoscopy cancellation/rescheduling), the unmatched cohort included 2,802 patients, with 253 (9.0%) receiving the educational video and 2,549 (91.0%) in the historical control group.

### Baseline Characteristics

For the primary outcome cohort, the intervention group had a significantly higher proportion of Latinx patients compared to the historical control group (47.2% vs 37.1%), while the control group had a higher proportion of Black patients (41.1% vs 33.5%).

After propensity score matching, baseline characteristics were well-balanced between the intervention and historical control groups (Table 1).

**Table 1:** Baseline Characteristics Primary Outcome Cohort Who Underwent. Colonoscopy Before and After Propensity Score Matching

|  | Unmatched |  |  | Matched |  |  |
| --- | --- | --- | --- | --- | --- | --- |
|  | Did not receive videos (N = 1471) | Received videos (N = 176) | P-value | Did not receive videos (N = 176) | Received videos (N = 176) | P-value |
| Age | 57.1 ± 8.1 | 56.2 ± 7.9 | 0.18 | 55.6 ± 7.3 | 56.2 ± 7.9 | 0.42 |
| Female, n (%) | 963 (65.5) | 116 (65.9) | 0.91 | 58 (33.1) | 60 (34.3) | 0.82 |
| Charlson Comorbidity Index | 2.2 ± 1.7 | 2.2 ± 1.6 | 0.80 | 2.0 ± 1.6 | 2.2 ± 1.6 | 0.23 |
| Race, n (%) |  |  | 0.03 |  |  | 0.96 |
| White | 105 (7.1) | 7 (4.0) |  | 9 (5.1) | 7 (4.0) |  |
| Black | 604 (41.1) | 59 (33.5) |  | 60 (34.3) | 59 (33.7) |  |
| Latinx | 546 (37.1) | 83 (47.2) |  | 79 (45.1) | 82 (46.9) |  |
| Other/Unknown | 216 (14.7) | 27 (15.3) |  | 27 (15.4) | 27 (15.4) |  |
| Insurance, n (%) |  |  | 0.10 |  |  | 0.80 |
| Medicaid | 431 (29.3) | 62 (35.2) |  | 60 (34.3) | 62 (35.4) |  |
| Medicare | 285 (19.4) | 20 (11.4) |  | 15 (8.6) | 20 (11.4) |  |
| Private | 724 (49.2) | 91 (51.7) |  | 97 (55.4) | 90 (51.4) |  |
| Self-pay | 30 (2.0) | 3 (1.7) |  | 3 (1.7) | 3 (1.7) |  |
| Workers' Comp | 1 (0.1) | 0 (0.0) |  | - | - |  |
| Language, n (%) |  |  | 0.93 |  |  | 0.71 |
| English | 1108 (75.3) | 132 (75.0) |  | 135 (77.1) | 132 (75.4) |  |
| Non-English | 363 (24.7) | 44 (25.0) |  | 40 (22.9) | 43 (24.6) |  |

For the secondary outcome cohort, the intervention group had a higher proportion of Latinx patients (47.4% vs 39.4%) and a lower proportion of Black patients (30.8% vs 39.6%) compared to the historical control group (p=0.032). There was also a trend toward a higher proportion of Medicaid patients in the intervention group (39.1% vs 31.5%), although the overall insurance distribution did not differ significantly between groups (p=0.062). After propensity score matching, baseline characteristics were well-balanced between the intervention and historical control groups (Table 2).

**Table 2:**
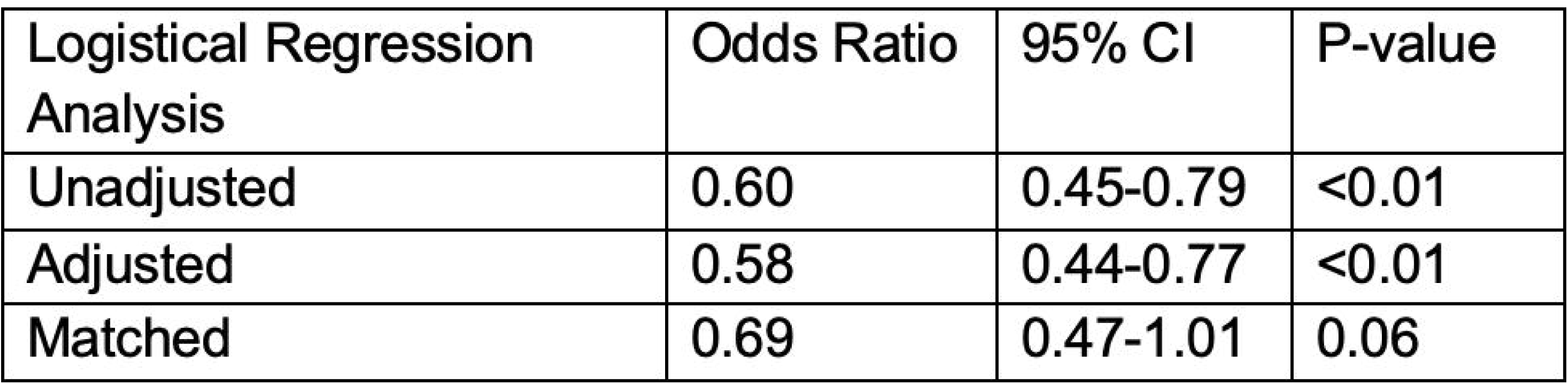
Baseline Characteristics Secondary Outcome Cohort Scheduled for Colonoscopy Before and After Propensity Score Matching.

To address potential confounding, propensity score matching was performed. After matching, baseline characteristics were well-balanced between the video and control groups (Table 1, Table 2). The standardized mean differences for all covariates were below 0.1, indicating adequate balance. The post-matching racial/ethnic distribution was similar between the two groups (p=0.957), as were the distributions of age, sex, Charlson comorbidity index, insurance type, and primary language (all p-values > 0.05).

### Primary Outcome: Inadequate Bowel Preparation

In the historical control group, the overall mean total BBPS score was 4.6 for colonoscopies with inadequate bowel preparation and 7.9 for adequate bowel preparation. In the intervention group, the overall mean total BBPS score was 3.6 for colonoscopies with inadequate bowel preparation and 7.8 for adequate bowel preparation. In the propensity score matched analysis of the primary outcome cohort, inadequate bowel preparation occurred in 12.0% of the intervention group versus 20.0% of the historical control group (p=0.041). Conditional logistic regression estimated a 48% reduction in odds of inadequate prep associated with the educational video (OR 0.52, 95% CI 0.28-0.96, p=0.038) (Table 3).

**Table 3:**
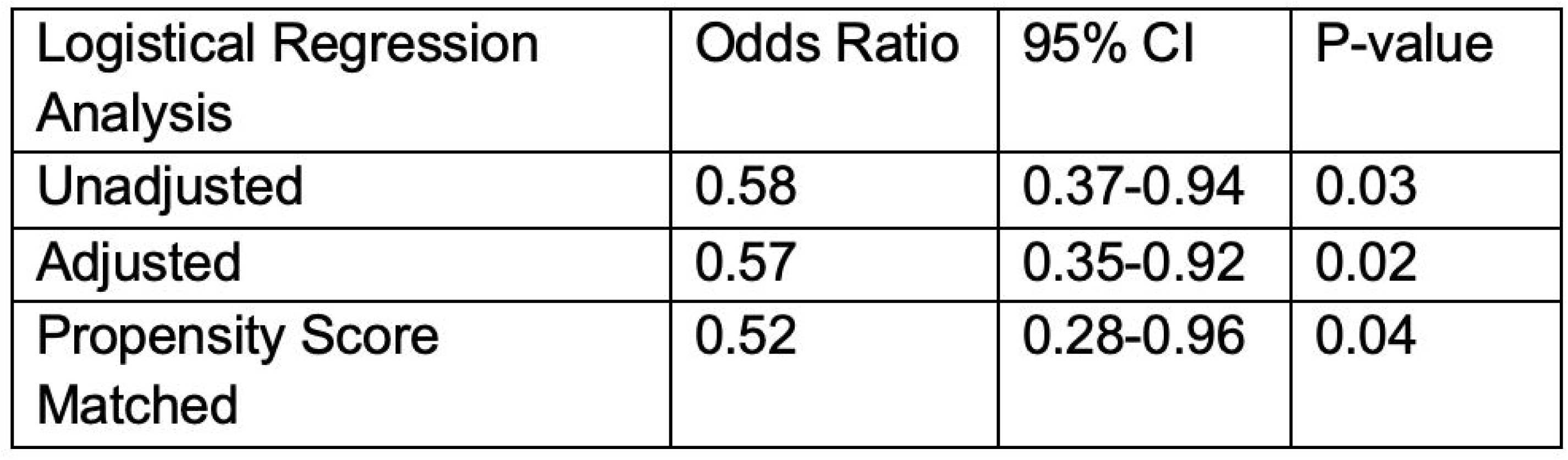
Odds of Inadequate Bowel Preparation Among Primary Outcome Cohort Who Underwent Colonoscopy.

The logistical regression model revealed similar findings. Inadequate bowel preparation occurred in 12.0% of patients in the intervention group compared to 18.8% in the historical control group (p=0.026). Unadjusted logistic regression showed that exposure to the educational video was associated with a 42% reduction in odds of inadequate prep (OR 0.58, 95% CI 0.37-0.94, p=0.028). After adjusting for potential confounders, the video remained associated with a 43% reduction in odds of inadequate prep (adjusted OR 0.57, 95% CI 0.35-0.92, p=0.022) (Table 3).

### Secondary Outcome: Colonoscopy Cancellations/Rescheduling

The propensity score matched analysis of the secondary outcome cohort showed a trend toward a 31% reduction in odds of cancellation or rescheduling associated with the educational video, although this did not reach statistical significance (OR 0.69, 95% CI 0.47-1.01, p=0.056) (Table 4).

**Table 4:** Odds of Colonoscopy Cancellation/Rescheduling Among Secondary Outcome Cohort Scheduled for Colonoscopy.

|  | Unmatched |  |  | Matched |  |  |
| --- | --- | --- | --- | --- | --- | --- |
|  | Did not receive videos (N = 2549) | Received videos (N = 253) | P-value | Did not receive videos (N = 253) | Received videos (N = 253) | P-value |
| Age | 56.8 ± 8.0 | 55.9 ± 8.1 | 0.09 | 56.4 ± 8.0 | 55.9 ± 8.1 | 0.52 |
| Female, n (%) | 925 (36.3) | 81 (32.0) | 0.18 | 77 (30.4) | 81 (32.0) | 0.70 |
| Charlson Comorbidity Index | 2.2 ± 1.8 | 2.2 ± 1.7 | 0.66 | 2.2 ± 1.9 | 2.2 ± 1.7 | 0.95 |
| Race, n (%) |  |  | 0.03 |  |  | 0.47 |
| White | 160 (6.3) | 14 (5.5) |  | 10 (4.0) | 14 (5.5) |  |
| Black | 1009 (39.6) | 78 (30.8) |  | 82 (32.4) | 78 (30.8) |  |
| Latinx | 1003 (39.4) | 120 (47.4) |  | 130 (51.4) | 120 (47.4) |  |
| Other/Unknown | 377 (14.8) | 41 (16.2) |  | 31 (12.3) | 41 (16.2) |  |
| Insurance, n (%) |  |  | 0.06 |  |  | 0.67 |
| Medicaid | 803 (31.5) | 99 (39.1) |  | 100 (39.5) | 99 (39.1) |  |
| Medicare | 480 (18.8) | 33 (13.0) |  | 37 (14.6) | 33 (13.0) |  |
| Private | 1200 (47.1) | 116 (45.9) |  | 114 (45.1) | 116 (45.9) |  |
| Self-pay | 63 (2.5) | 5 (2.0) |  | 2 (0.8) | 5 (2.0) |  |
| Workers' Comp | 3 (0.1) | 0 (0.0) |  | - | - |  |
| Language, n (%) |  |  | 0.25 |  |  | 1.00 |
| English | 1928 (75.6) | 183 (72.3) |  | 183 (72.3) | 183 (72.3) |  |
| Non-English | 621 (24.4) | 70 (27.7) |  | 70 (27.7) | 70 (27.7) |  |

For the logistical regression model, 30.4% of patients who received the educational video had a cancelled or rescheduled procedure compared to 42.3% of controls. This difference was statistically significant (p<0.001). Unadjusted logistic regression found that the video was associated with a 40% reduction in odds of cancellation or rescheduling (OR 0.60, 95% CI 0.45-0.79, p<0.001). After adjusting for potential confounders, the video remained associated with a 42% reduction in odds of cancellation or rescheduling (adjusted OR 0.58, 95% CI 0.44-0.77, p<0.001) (Table 4).

### Subgroup Analyses

Subgroup analyses were performed to assess the impact of the educational video on the primary outcome of inadequate bowel preparation stratified by sex, race/ethnicity, Charlson comorbidity index, insurance status, and primary language (Supplementary Table 1). In the unmatched cohort, the video was associated with a significant reduction in inadequate prep among females (OR 0.52, 95% CI 0.27-0.99, p=0.046) and patients whose primary language was English (OR 0.53, 95% CI 0.30-0.94, p=0.031). The video was not associated with a significant reduction in adequate prep amongst males (OR 0.69, 95% CI 0.34-1.41, p=0.308) and patients whole primary language was not English (OR 0.75, 95% CI 0.32-1.74, p=0.500). These findings suggest that the educational video may be particularly beneficial for these subgroups. Trends toward a reduction in inadequate prep were observed in other subgroups, including males, English speakers, and those with Charlson comorbidity index <3, although these findings did not reach statistical significance.

## Discussion

In this single-center, observational study, we found that an educational video intervention was associated with a large and significant reduction in inadequate bowel preparation among patients who underwent colonoscopy. In addition, the educational video intervention was associated with a large and significant reduction in cancellation or rescheduling of colonoscopy. Both findings remained after adjustment for potential confounders and in propensity score-matched analyses. These findings suggest that an educational video may be an effective tool for optimizing patient preparation for colonoscopy, a critical factor in ensuring high-quality examinations and early detection of colorectal cancer.

Previous studies on the impact of educational interventions on bowel preparation quality have reported mixed findings. A randomized controlled trial by Calderwood et al. found that a brief educational video improved bowel preparation quality compared to standard instructions, with a 76% adequacy rate in the video group versus 46% in the control group (p=0.003).^11^ However, a study by Veldhuijzen et al. found no significant difference in bowel preparation quality between patients who received an educational video and those who received standard instructions (p=0.43).^12^

Our study demonstrates that a short, automated video-based intervention can significantly improve bowel preparation quality and reduce colonoscopy cancellations and rescheduling. The use of a 90-second video delivered via text messaging represents a novel, scalable approach that addresses limitations of previous studies, such as the need for longer videos or more labor-intensive delivery methods. The automated nature of our intervention makes it easily able to be disseminated to large patient populations without requiring significant additional resources, highlighting its potential for widespread implementation in clinical practice.

Notably, our educational video not only improved bowel preparation quality but also reduced colonoscopy cancellations and rescheduling. This finding suggests that the video may have addressed factors beyond bowel preparation instructions, such as emphasizing the importance of the procedure and allaying patient concerns. The combined impact of improved bowel preparation and reduced cancellations has the potential to substantially enhance the efficiency and effectiveness of colonoscopy screening programs.

Furthermore, the principles of our intervention - using short, engaging videos delivered via automated text messaging - could be adapted to improve patient education and adherence in other healthcare contexts. Similar approaches could be used to prepare patients for other medical procedures, support medication adherence, or encourage lifestyle changes for chronic disease management. Our findings highlight the broad potential of video-based interventions to positively impact patient care across various settings.

Our study has several limitations that should be acknowledged. First, as a single-center study, our findings may not be generalizable to other patient populations or healthcare settings. Second, the observational, non-randomized design of our study may introduce selection bias and confounding, although we attempted to mitigate these issues through multivariable adjustment and propensity score matching. Despite these limitations, our study has important implications for clinical practice and future research. Our findings suggest that an educational video intervention may be a simple, scalable strategy for improving bowel preparation quality and reducing colonoscopy cancellations and rescheduling. Future studies should investigate the implementation and effectiveness of educational videos in diverse healthcare settings, as well as explore the potential for tailoring these interventions to specific patient subgroups. Randomized controlled trials with larger sample sizes and longer follow-up periods may help to establish the causal relationship between educational videos and improved colonoscopy outcomes.

We acknowledge that other changes in practice could have occurred during the study period. However, to the best of our knowledge, no other significant changes in pre-colonoscopy protocols or patient communication were implemented during this time. The endoscopy unit maintained consistent staffing and scheduling practices throughout the study period. We acknowledge that the time frame of our historical control group was still within the Covid-19 epidemic and that this could have affected patient’s behaviors including increased concerns about proceeding with procedures and lower thresholds to cancelling or rescheduling. However, during this time frame, our institution no longer required pre-procedural Covid-19 testing. Future studies should investigate metrics such as insertion time, total procedure time, cecal intubation rate, or adenoma detection rate on quality of bowel preparation. Additionally, we were unable to adjust for BMI in our analysis due to the lack of reliable BMI data at the time of the procedures. It’s worth noting that existing literature shows conflicting evidence regarding the impact of BMI on colonoscopy bowel preparation.^13,14,15^ While segmental BBPS scores were not consistently available in our colonoscopy reports, we relied on total BBPS scores which have been previously validated as a reliable measure of bowel preparation adequacy.^16^

In conclusion, we found that an educational video intervention was associated with significant improvements in bowel preparation quality and a reduction in colonoscopy cancellations and rescheduling in a diverse, real-world patient population. Our findings highlight the potential value of educational videos as a simple, scalable strategy for optimizing patient preparation for colonoscopy and improving the quality and efficiency of this important screening and diagnostic tool. Further research is needed to validate these findings in other healthcare settings and to explore the potential for tailoring educational interventions to specific patient subgroups.

## Data Availability

All data produced in the present work are contained in the manuscript.

## Figures/Tables

Supplementary Table 1: Results of Subgroup Analysis

