## Supplemental Table 1 for "Impact of a Smart Phone-Based Educational Video on Bowel Preparation and Colonoscopy Adherence: A Propensity Score Matched Analysis"

| Subgroup Analysis | Odds Ratio | 95% CI | P-value |
|---|---|---|---|
| Sex | | | |
| Female | 0.52 | 0.27-0.99 | 0.05 |
| Male | 0.69 | 0.34-1.41 | 0.31 |
| Language | | | |
| English | 0.53 | 0.30-0.94 | 0.03 |
| Non-English | 0.75 | 0.32-1.74 | 0.50 |
| Charlson Comorbidity Index | | | |
| Score < 3 | 0.55 | 0.30-0.99 | 0.05 |
| Score >= 3 | 0.67 | 0.31-1.45 | 0.31 |
| Insurance | | | |
| Medicaid | 0.58 | 0.27-1.21 | 0.15 |
| Medicare | 0.43 | 0.10-1.93 | 0.27 |
| Private | 0.64 | 0.32-1.27 | 0.20 |
| Self-pay | 1.00 | - | - |
| Race | | | |
| White | 1.00 | - | - |
| Black | 0.41 | 0.16-1.04 | 0.06 |
| Latinx | 0.85 | 0.47-1.54 | 0.58 |
| Other/Unknown | 0.19 | 0.03-1.46 | 0.11 |
